# Specific Within-Domain Cognitive Impairments Predict Depression Severity Six-Months After Stroke

**DOI:** 10.1101/2024.09.25.24314204

**Authors:** Kyle Kelleher, Nele Demeyere, Andrea Kusec

## Abstract

**Background:** Following stroke, chronic cognitive impairments across multiple domains have been associated with depression. Currently, it is unknown if specific subtypes of cognitive impairments differentially relate to post-stroke depression severity. This study aimed to explore the differential associations between within-domain cognitive impairment to depression severity six-months after stroke.

**Method:** Participants (*n* = 385, Age *M* = 73.86 years [*SD* = 12.51], National Institutes of Health Stroke Severity *M* = 6.83 [*SD* = 6.01]) were recruited from an acute stroke ward. Participants completed a self-report mood measure (Hospital Anxiety and Depression Scale; HADS) and a stroke-specific cognitive assessment (Oxford Cognitive Screen; OCS). Separate multiple regressions predicting depression were conducted across 1) OCS domain-specific cognitive impairments of language, memory, attention, praxis, numeracy and executive function, and 2) the novel subtask-specific impairments within each OCS domain. Anxiety severity and years of education attained were included as covariates.

**Results:** Within-domain impairments that were uniquely associated with depression severity were calculation (*b*_(.57)_ = 1.44, 95% *CI* [0.31, 2.56], *p* = .012), episodic memory (*b*_(.52)_ = 1.36, 95% *CI* [0.34, 2.37], *p* = .009), picture naming (*b*_(.45)_ = 1.18, 95% *CI* [0.31, 2.06], *p* = .008), number writing (*b*_(.46)_ = 2.54, 95% *CI* [0.26, 2.07], *p* = .012), and visuospatial attention (*b*_(.35)_ = 1.24, 95% *CI* [0.54, 1.93], *p* = .001). Analysis in pooled multiply imputed data (*N* = 430) corroborated complete case analysis findings.

**Conclusions:** Specific within-domain cognitive impairments have differential relationships with post-stroke depressive symptomology. Accommodating for these impairments in post-stroke depression interventions may potentially enhance therapeutic outcomes.

## Introduction

Stroke is a leading cause of disability^1^ which frequently results in physical impairments, such as hemiparesis, as well as cognitive impairments, including poor memory and attention.^2^ Cognitive impairment is particularly common, with prevalence rates ranging from 34–60% long term after stroke.^3,4^

One in three stroke survivors will experience depression following stroke,^5^ characterised by a marked decrease in mood and/or a loss of interest in daily activities.^6^ Post-stroke depression has been shown to negatively impact neurorehabilitation and reduce long-term gains.^7,8^ Crucially, post-stroke depression has been repeatedly associated with general cognitive impairment in systematic reviews.^9,10^ Greater severity of cognitive impairment still correlates with lower quality of life even beyond 2-years post-stroke,^4^ suggesting a persistent mood-cognition link over time.

Research investigating the mood-cognition link has largely been examined using screening measures of domain-general cognition such as the Montreal Cognitive Assessment (e.g., Vermeer et al.^11^). However, post-stroke cognitive impairment can occur within single or multiple overlapping but distinct areas of cognition (e.g., memory, attention). These can be conceptualised as ‘domain-specific’ impairments that may have differential impacts on post-stroke function.^12,13^ Of the domain-specific relationships that have been investigated in neurological populations, unique links have been evidenced between depression to verbal memory, working memory, and executive function.^14^ More recently, investigations into across-domain cognitive impairments have delineated a more granular mood-cognition relationship in stroke survivors. Williams & Demeyere^15^ found that memory, language, spatial attention, executive function, numeracy, and praxis impairments all predicted increases in depression severity, even after covarying for co-occurring anxiety. Importantly, the association between these domains and depression had differential strengths (β range = 1.18 [memory] – 2.06 [praxis]). Notably, there was no significant association between cognitive impairment and anxiety severity six-months after stroke after covarying for co-occurring depression severity.^15^ This suggests cognitive impairment uniquely relates to post-stroke depression but not anxiety severity in this population, and that the strength of this association may vary depending on the domain of cognition impaired.

However, domain-specific impairments are composed of several within-domain “subtypes”. For example, post-stroke language impairments could occur purely in terms of difficulties with semantic understanding, or as difficulties with speech production. To our knowledge, there are no investigations of the relationship of within-domain cognitive impairments to post-stroke depression. Stroke survivors experiencing an impairment in one broad domain of cognition, such as memory, may face challenges for various reasons, each with distinct implications for their care and functioning. It has been shown that within-domain cognitive impairments can occur independent of one another.^16^ More precise information on which specific within-domain impairments most strongly link with depression could yield important information on how post-stroke depression is maintained and in developing more targeted intervention techniques.

Whereas Williams and Demeyere (2021)^15^ explored a population with relatively mild stroke severity (NIHSS <3 = 65.9%), there is benefit in examining the mood-cognition relationship in a more moderately impaired stroke sample. Possibly, differential relationships emerge in mild versus more moderately impaired stroke samples, which is yet unexamined.

In this study, we aimed to both replicate the association between domain-specific cognitive impairment to post-stroke depression as in Williams & Demeyere^15^ in a separate stroke cohort, and 2) extend this investigation by exploring how within-domain cognitive impairments contribute to post-stroke depression using a more cognitively impaired sample.

## Methods

### Study Design

This study presents a retrospective analysis of a cross-sectional observational within-subjects design. The STROBE guideline was used for reporting. Data is freely available at https://www.dementiasplatform.uk/ and analysis code at https://osf.io/yrth4/

### Sample

#### Eligibility and Recruitment Procedure

Data was retrospectively analysed from the multi-site Oxford Cognitive Screening Programme^13^ (National Research Ethics Committees REC reference 18/SC/0550). Eligibility criteria were:

1. Age ≥18 years.
2. Medical diagnosis of stroke.
3. Capacity to provide informed consent.
4. Ability to concentrate for 15 – 20 minutes.

Recruitment occurred in the Oxford John Radcliffe Hospital acute stroke ward between 2012 – 2020. Participants consenting to the Oxford Cognitive Screening programme additionally opted to complete a six-month post-stroke assessment of mood and cognition, forming the present study data.

### Study Measures

Stroke characteristics, including sex, years of education, age at time of stroke, type of stroke (ischaemic vs haemorrhagic), and lesion hemisphere were obtained from medical records with participant consent. Acute stroke severity was assessed using the clinician-rated NIHSS^17^ from 0-42 points where higher scores indicate higher stroke severity. Depression and anxiety severity were measured on the HADS depression (HADS-D) and anxiety (HADS-A) subscales, respectively.^18^ Higher scores indicate worse mood. Scores greater than 7 on either subscale indicate clinically significant anxiety/depression. Both the depression (Cronbach’s α = .76) and anxiety (α = .87) subscales show good internal consistency six-months after stroke.^19^

The Oxford Cognitive Screen (OCS)^13^ measures six cognitive domains (with 11 subtasks) to screen for cognitive impairments commonly caused by stroke: language (picture naming, semantics, sentence reading), numeracy (number writing, calculation), praxis (gesture imitation), memory (orientation, semantic memory, episodic recognition memory), visuospatial attention (sustained visual attention, egocentric neglect, allocentric neglect), and executive function (trail-making task).

Subtask responses are binarised indicating impairment (1) or no impairment (0) based on published cut-off scores from a normative sample.^13^ Impairment on any of the subtasks within a domain constituted binary domain-specific impairment.

### Sample Size

#### Post-hoc Power Analysis

Given this study was a secondary analysis of existing data, sample size (*N* = 430) was determined prior to analysis. Using the ‘*pwr’* package^20^ in R, the complete case data (*N* = 385) has 99% power to detect an effect size of 0.25 with an alpha of 0.05 as a minimal effect size of interest.

#### Analysis Plan

All analyses were performed using ‘*Rstudio’* (version 2022.07.2) and ‘*R*’ (version 4.2.2: R Core Team, 2022). Descriptive statistics were carried out across all study variables.

Direct comparisons on severity of cognitive impairment, mood, and demographic variables between the present stroke sample and the separate study sample analysed in Williams & Demeyere^15^ were conducted to more precisely describe the sample differences (see *Supplementary Materials*).

A series of multiple linear regression analyses examined the relationship of 1) the *domain-specific* cognitive impairments, and 2) the *subtask-specific* cognitive impairments within each domain to post-stroke depression and anxiety severity. Unstandardized b-estimates are reported with 95% confidence intervals (CIs). The assumptions of multiple linear regression were tested for all models.

The high within-subject covariance between self-reported anxiety and depression^21^ can become conflated with the variance in the association between depression and cognitive impairment in stroke survivors^15^, spuriously increasing the strength of this latter relationship when measured. Therefore, the Williams and Demeyere^15^ two-model approach was replicated here: predicting depression severity with, and without covarying for anxiety severity. These models were labelled “Depression Only Models” and “Depression with Anxiety Models”, respectively, throughout. Reporting on, and directly comparing, both types of models, aids in untangling the association between cognitive impairment and depression severity from the confounding effect of anxiety severity, ultimately increasing confidence in the findings.

False discovery rate (FDR) corrections were applied in across-domain analyses to reduce Type-I error inflation when performing multiple tests.^22^ Any significant domain-specific predictor variables in the FDR-corrected regression models were explored at the subtask-specific level using the same analysis procedure.

#### Determining model covariates

Pairwise Pearson correlations were performed between depression and anxiety severity with age stroke, years of education, NIHSS scores, with a two-tailed independent *t-* test for determining an association between participant sex. Significantly related variables (alpha < .05) were included as covariates. Since the aim of the present study is to explore in greater depth an already identified association between domain-specific cognitive impairment and depression severity^15^ this data-driven approach to covariance was chosen to increase the predictive accuracy.^23^

#### Missingness of Data

Missingness was classified as participants with incomplete OCS data. Analysis was performed on both complete case data (*n* = 385 of 430) and on a dataset generated by pooling results from five multiply imputed datasets with a maximum of 50 iterations to maximise use of data and determine stability of findings. Imputations were conducted using predictive mean matching (PMM) using the ‘*mice’* R package.^24^

## Results

### Descriptive Statistics

Information about stroke type and demographic information and is presented in Table 1.

**Table 1:** Stroke and Demographic Information.

| Stroke Type | *n* (%) |
| --- | --- |
| Ischaemic | 255 (66.23) |
| Haemorrhagic | 62 (16.10) |
| Undetermined from scan | 65 (16.85) |
| Mixed | 3 (0.78) |
| Lesion Hemisphere |  |
| Right | 152 (39.49) |
| Left | 137 (35.58) |
| Bilateral | 66 (17.14) |
| Undetermined from scan | 30 (7.79) |
| Stroke Recurrence |  |
| First | 264 (68.37) |
| Recurrent | 121 (31.43) |
| Demographics |  |
| Age (Years) | 73.86 (12.51) |
| Education Attained (Years) | 12.25 (3.55) |
| NIHSS Score | 6.83 (6.01) |
| Male Sex (%) | 54.03 |

*Note.* This table summarises complete case data (*n* = 385), OCS = Oxford Cognitive Screen, NIHSS = National Institute for Health Stroke Severity

Full details of cohort comparisons are in the *Supplementary Materials.* In brief, the present cohort had significantly greater stroke severity and a significantly greater proportion of domain-specific cognitive impairments compared to the stroke cohort reported on by Williams and Demeyere.^15^

The assumptions of homoscedasticity, linearity, and non-multicollinearity were met for all regression models. However, the assumption of normality of residuals was only partially satisfied, for all models due to the left skewness of self-report mood data clustering around zero (see *Supplementary Materials*).

### Association of Cognitive Impairment with Depression

#### Depression Model Covariates

HADS-D only significantly correlated with HADS-A (*r*_(383)_ = .63 *p* < .001), and years of education (*r*_(380)_ = −.001 *p* = .044). Therefore, years of education attained (hereafter ‘education’) is included as a covariate for all models predicting depression severity. HADS-A is included as a covariate for the Depression with Anxiety Models only.

#### Replication - Domain-Specific Models Predicting Depression Severity

##### Depression Only Models

Without accounting for co-occurring anxiety severity, the domain-specific impairments that were significant predictor variables were language (*b*_1(0.46)_ = 1.24, 95% *CI* [0.33, 2.14], *t* = 2.70, *p* = .007), numeracy (*b*_1(0.53)_ = 1.86, 95% *CI* [0.82, 2.89], *t* = 3.52, *p* < .001), memory (*b*_1(0.46)_ = 1.35, 95% *CI* [0.45, 2.26], *t* = 2.93, *p* = .004), and spatial attention (*b*_1(0.42)_ = 1.27, 95% *CI* [0.44, 2.11], *t* = 3.00, *p* = .003). Neither praxis nor executive function were significant predictor variables in their respective models. Education was a non-significant predictor variable in all models.

Every Depression Only model was significant (*p* range = .002 [numeracy] – .021 [language]), except for those including praxis and executive function. Explained variance was small for all models (*Adjusted R*^2^ range = .02 [language] – .04 [numeracy], *Supplemental Materials*). Pooled imputation results aligned with complete case analysis (complete case *t*-values range = 1.56 [executive function] – 3.52 [numeracy]; pooled imputation *t*-values range = 1.50 [executive function] – 3.57 [numeracy]: *Supplementary Materials*).

##### Depression with Anxiety Models

After accounting for co-occurring anxiety severity, domain-specific impairments that continued to be associated with depression severity were language (*b*_1(0.36)_ = 0.76, 95% *CI* [0.05, 1.57], *t* = 2.11, *p* = .036), numeracy (*b*_1(0.42)_ = 1.28, 95% *CI* [0.46, 2.10], *t* = 3.01, *p* = .002), memory (*b*_1(0.36)_ = 0.91, 95% *CI* [0.20, 1.63], *t* = 2.53, *p* = .012), spatial attention (*b*_1(0.33)_ = 0.85, 95% *CI* [0.19, 1.50], *t* = 2.54, *p* = .012). Praxis now additionally predicted depression severity (*b*_1(0.42)_ = 0.94, 95% *CI* [0.89, 3.48], *t* = 2.27, *p* = .024), but not executive function. Anxiety severity, but not education, was a significant predictor in all models (*ps <* .001).

Every Depression with Anxiety model was significant (*ps* < .001, *Adjusted R*^2^ range = .38 [executive function] – .40 [numeracy]). Pooled imputation results aligned with complete case analysis (complete case *t*-values range = 1.82 [executive function] – 3.01 [numeracy]; pooled imputation *t*-values range = 2.00 [executive function] – 3.31 [numeracy]: *Supplemental Material*).

#### Extension - Subtask-Specific Models Predicting Depression Severity

##### Depression Only Models

Without accounting for co-occurring anxiety severity, the only subtask-specific impairments that were significant predictor variables in models that significantly predicted depression severity were calculation (*p* < .001), episodic memory (*p* = .002), and visuospatial attention (*p* < .001: Table 2). Education was a non-significant predictor in all models. Praxis is a domain comprising of one test on the OCS, so no further subtask-specific models were performed.

**Table 2:** ‘Depression Only Models’ Predicting Depression Severity by Subtask-Specific OCS Cognitive Impairments and Education.

| Depression Severity $\sim b_0 + b_1$ Impairment + $b_2$ Education + $\epsilon$ | | | | | | | |
| --- | --- | --- | --- | --- | --- | --- | --- |
| Impairment | Overall Model Statistics | | | $b_1$ Impairment Statistics | | | |
| | *F* statistic (*df*) | *AdjR*<sup>2</sup> | *p* value | $b_1$ (*SE*) | *t* statistic | 95% *CI* | *p* value |
| Picture Naming | 3.78 (2, 378) | .01 | .260 | 1.09 (0.57) | 1.91 | [-0.03, 2.22] | .057 |
| Semantic Task | 4.31 (2, 227) | .03 | .160 | 3.18 (1.25) | 2.53 | [0.70, 5.65] | .012* |
| Sentence Reading | 3.38 (2, 376) | .01 | .385 | 0.88 (0.54) | 1.64 | [-0.17, 1.93] | .101 |
| Number Writing | 5.41 (2, 373) | .02 | .053 | 1.58 (0.58) | 2.71 | [0.43, 2.73] | .007* |
| Calculation | 7.73 (2, 372) | .03 | .006* | 2.48 (0.72) | 3.44 | [1.06, 3.89] | < .001* |
| Orientation | 2.67 (2, 377) | .01 | .775 | 0.71 (0.58) | 1.23 | [-0.43, 1.86] | .218 |
| Verbal Memory | 3.04 (2, 370) | .01 | .538 | 0.77 (0.56) | 1.38 | [-0.33, 1.87] | .168 |
| Episodic Memory | 6.96 (2, 370) | .03 | .012* | 2.03 (0.65) | 3.11 | [0.75, 3.31] | .002* |
| Broken Heart Task | 8.06 (2, 372) | .04 | .004* | 1.52 (0.45) | 3.39 | [0.64, 2.40] | < .001* |
| Space Attention | 2.46 (2, 368) | .01 | .956 | 0.54 (0.64) | 0.85 | [-0.71, 1.79] | .397 |
| Object Attention | 2.50 (2, 368) | .01 | .922 | 0.45 (0.50) | 0.89 | [-0.54, 1.43] | .374 |

*Note.* This table summarises 11 univariate models predicting depression severity with 11 of the novel OCS subtask impairments where the domain-level impairments were significant, covarying for education. ‘Broken Hearts’ is a task indexing visuospatial attention. ‘Space’ and ‘Object Attention’ are tasks indexing egocentric and allocentric neglect, respectively. \*Significant ( $p < .050$ ) after false discovery rate correction for multiple comparisons.

Pooled imputation results aligned with all complete case analyses (complete case *t*-values range = .85 [object attention] – 3.39 [visuospatial attention]; pooled imputation *t*-values range = .68 [space attention] – 3.37[visuospatial attention]: *Supplemental Material*).

##### Depression with Anxiety Models

After accounting for co-occurring anxiety severity, picture naming (*p* = .008), number writing (*p* = .011), calculation (*p* = .012), episodic recognition (*p* = .009), and visuospatial attention (*p* < .001: Table 3) still predicted depression severity. Anxiety severity, but not education, was a significant predictor variable (*ps <* .001). All Depression with Anxiety Models were significant (*p*s < .001).

**Table 3:** ‘Depression with Anxiety Models’ Predicting Depression Severity by Subtask-Specific OCS Cognitive Impairment, Education, and Anxiety Severity.

| Impairment | Depression Severity $\sim b_0 + b_1$ Impairment + $b_2$ Education + $b_3$ Anxiety Severity + $\epsilon$ | | | | | | |
| --- | --- | --- | --- | --- | --- | --- | --- |
| | Overall Model Statistics | | | $b_1$ Impairment Statistics | | | |
| | *F* statistic (*df*) | *AdjR*<sup>2</sup> | *p* value | $b_1$ (*SE*) | *t* statistic | 95% *CI* | *p* value |
| Picture Naming | 86.65 (3, 377) | .40 | < .001* | 1.18 (0.45) | 2.66 | [0.31, 2.06] | .008* |
| Semantic Task | 48.16 (3, 226) | .38 | < .001* | 1.37(1.01) | 1.35 | [-0.63, 3.36] | .179 |
| Sentence Reading | 82.39 (3, 375) | .39 | < .001* | 0.47 (0.42) | 1.13 | [-0.35, 1.30] | .260 |
| Number Writing | 85.16 (3, 372) | .40 | < .001* | 1.17 (0.46) | 2.54 | [0.26, 2.07] | .011* |
| Calculation | 83.91 (3, 371) | .40 | < .001* | 1.44 (0.57) | 2.51 | [0.31, 2.56] | .012* |
| Orientation | 85.24 (3, 376) | .40 | < .001* | 0.75 (0.45) | 1.66 | [-0.14, 1.64] | .098 |
| Verbal Memory | 80.12 (3, 369) | .39 | < .001* | 0.56 (0.44) | 1.28 | [-0.30, 1.42] | .203 |
| Episodic Memory | 83.02 (3, 369) | .40 | < .001* | 1.36 (0.52) | 2.63 | [0.34, 2.37] | .009* |
| Broken Hearts Task | 86.05 (3, 371) | .41 | < .001* | 1.24 (0.35) | 3.50 | [0.54, 1.93] | <.001* |
| Space Attention | 80.36 (3, 367) | .39 | < .001* | 0.75 (0.50) | 1.51 | [-0.23, 1.73] | .132 |
| Object Attention | 79.13 (3, 367) | .41 | < .001* | 0.14 (0.38) | 0.20 | [-0.70, 0.86] | .716 |

*Note.* This table summarises 11 models predicting depression severity by 11 of the novel OCS subtask impairments where domain-level impairments were significant, covarying for anxiety and education. ‘Broken Hearts’ indexes visuospatial attention. ‘Space’ and ‘Object Attention’ index egocentric and allocentric neglect, respectively. \*Significant ( $p < .050$ ) after false discovery rate correction for multiple comparisons.

Pooled imputation results aligned with all complete case analyses (complete case *t*-values range = .20 [object attention] – 3.50 [visuospatial attention]; pooled imputation *t*-values range = .36 [object attention] – 3.37 [visuospatial attention]: *Supplementary Materials*).

### Summary of Replication Results and Novel Depression Severity Results

The findings of Williams and Demeyere (2021)^15^ study were replicated here in a more moderately impaired stroke cohort. The proportion of concurrent domain-specific impairments, or ‘domain-general’ impairment, significantly predicted depression severity before and after covarying for co-occurring anxiety severity. Additionally, in line with Williams & Demeyere (2021)^15^ neither domain-general impairment nor any domain-specific cognitive impairment predicted anxiety severity after co-occurring for depression severity (*see Supplemental Material*).

In our novel within-domain analyses, only five of the novel 11 tested OCS subtask-specific cognitive impairments significantly predicted depression severity after accounting for co-occurring anxiety severity and years of education attained (Figure 1). These tasks were calculation, episodic recognition, spatial attention accuracy, picture naming, and number writing. Further, each subtask-specific cognitive impairment associated with depression with differential strengths – with performance on the calculation task demonstrating the strongest link (Figure 2).

**Figure 1:**
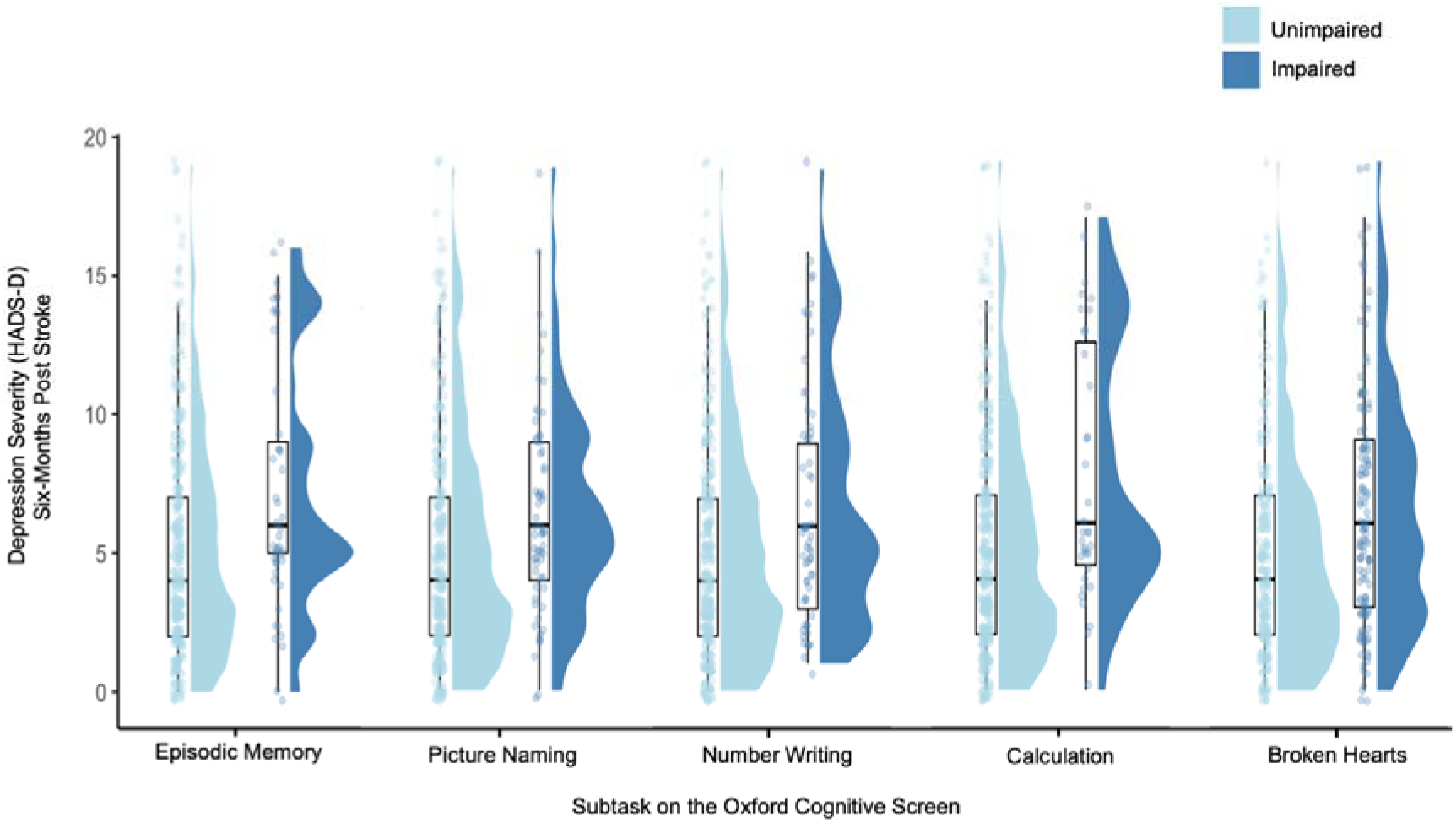
Raincloud Plots of Six-Month Post-Stroke Depression Severity by Cognitive Impairment Note. HADS-D = Hospital Anxiety and Depression Scale-Depression

**Figure 2:**
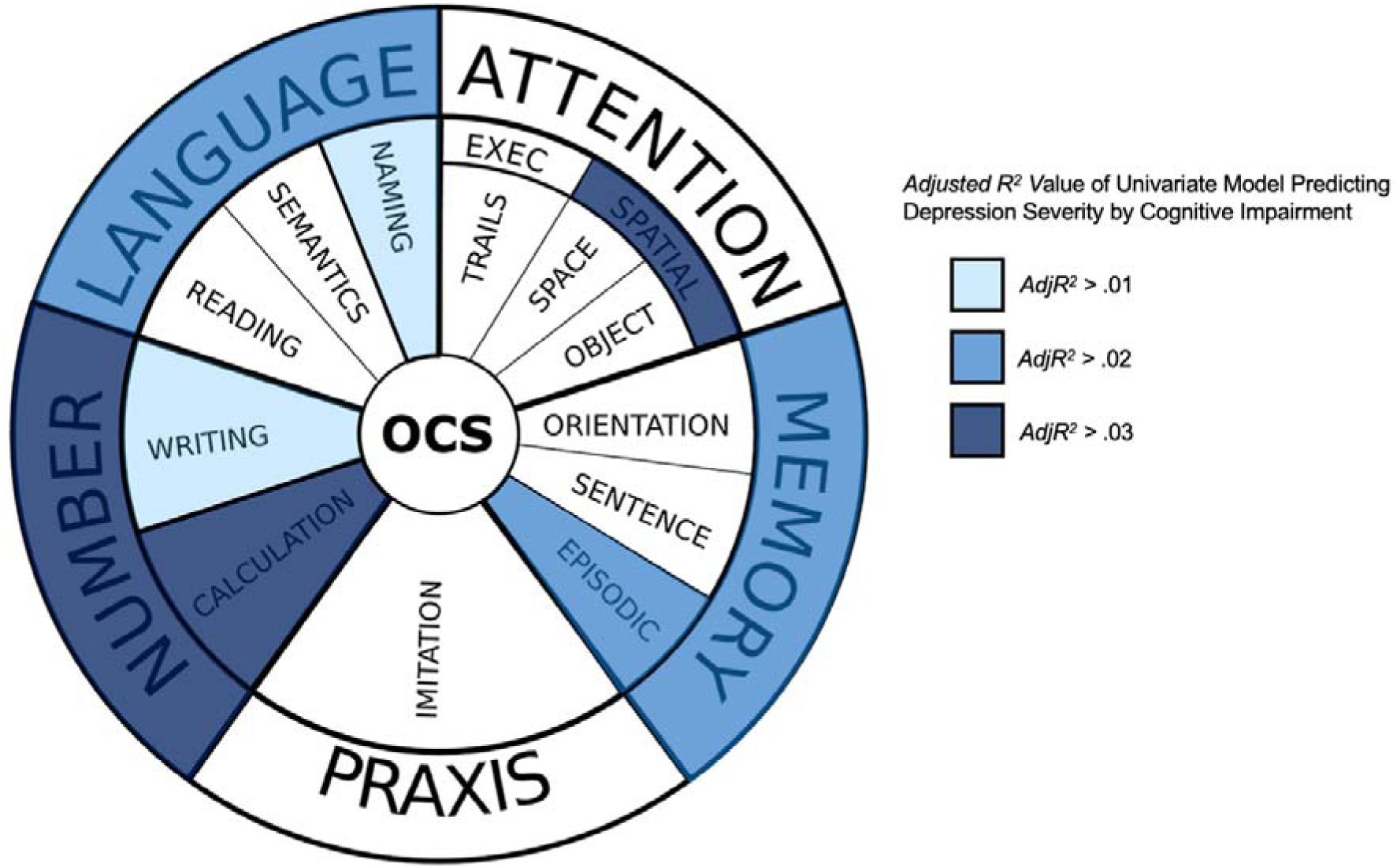
Heatmap of the Domain- and Subtask-Specific Impairments Predictive of Depression Severity Six-Months After stroke

## Discussion

### Domain-Specific Impairments

This study explored the association between cognitive impairments to depression severity six-months after stroke. This replication and extension of the Williams and Demeyere^15^ study was performed in a stroke sample with more frequent, and severe, cognitive impairments, while detailing the contribution of within-domain cognitive impairments to depression severity. As in Williams and Demeyere^15^ impairments in memory, language, numeracy, spatial attention, and praxis domains predicted depression severity when accounting for co-occurring anxiety severity. Similarly, each association had differential strengths, suggesting that the effects of post-stroke cognitive impairment on depression severity are heterogenous. Exploring the link between post-stroke mood and cognition with highly granular cognitive outcomes aligns with national guidelines to conceptualise cognitive impairment as multifactorial for post-stroke rehabilitation.^25,26^

In contrast to Williams and Demeyere^15^ domain-level executive function impairment was not found to be associated with 6 months depression severity in this sample. It is possible that there is a lower perceivable impact of executive function on everyday activities in the context of more severe stroke. For instance, a study of older adults with neurocognitive disorders found that executive function impairment negatively correlated with performance on complex, multistep activities of daily living (ADLs) but not functional tasks like washing and dressing oneself.^27^ Therefore, the effects of this impairment may decrease in perceptual salience as stroke severity increases, especially considering that survivors of more severe strokes are more likely to have assistance with complex ADLs.^28^ Additionally, the OCS measures executive function via only one task switching assessment. It is possible that other aspects of executive functions note measured in a simple trails task could potentially contribute to depression after stroke. It is worth noting in this context that the OCS calculation task may have inadvertently indexed an association between working memory impairment and depression severity. Finally, the role of effort in executive tasks should be considered, given that depression more generally is associated with reduced effort and increased mental fatigue in demanding cognitive tasks.^4,29^ The OCS task switching assessment using circles and squares instead of letters and numbers as a means of reducing verbal effort for those with aphasia; however, this possibly alters the effort required for stroke survivors without aphasia. This opens up avenues for future research to investigate a conceivable relationship linking effort, post-stroke depression, and different sub-types of executive function.

Anxiety and education were included as covariates in these models. Despite the strong association of anxiety with depression in stroke,^30^ cognitive impairment still had a greater impact of change on depression severity than anxiety severity – indicated by greater *b* coefficients in all models. This suggests that post-stroke cognitive impairment is most strongly associated with depression severity.

### Within-Domain Cognitive Impairments and Depression Severity

In addition to the replication of Williams and Demeyere^15^ are the novel associations identified between performance on specific within-domain tasks on the OCS and depression severity. Impairments in episodic memory, calculation, number writing, picture naming, and visuospatial attention tasks were all uniquely associated with increased depression severity – even after accounting for co-occurring anxiety severity.

The potential mechanisms underlying these associations are likely dynamic and manifold. In terms of memory, increased severity of depressive symptoms has been associated with reduced episodic memory in both stroke^31^ and non-stroke populations.^32^ Therefore, an acquired episodic recognition impairment could exacerbate and maintain existing symptoms of low mood, in turn worsening episodic memory, mutually perpetuating one another. Impaired episodic memory can also hinder even basic aspects of socialising like remembering what was said in a conversation, which has been documented to increase social isolation and frustration in other neurological populations.^33^

Stroke survivors identify memory difficulties as a long-term issue in patient reports of unmet needs, negatively impacting quality of life.^34,35^ Memory impairments have a tangible impact in daily life such as remembering how and when to execute daily activities and leisure activities, and remembering having completed them.^36^ This enhanced perceptual salience of memory impairment could generate emotional distress in response to its impact.

With regards to the number domain, mental arithmetic requires resources from working memory which is thought to mediate the maintenance of depression outside of stroke.^37^ Likewise, the observed association between post-stroke impairments in the number writing task and depression severity may also be indicative of this potential connection to working memory. Alternatively, impaired calculation ability may hinder engagement in ADLs like managing finances, thereby reducing autonomous participation in society^38^ and worsening mood. Similarly, the number writing task involves transcribing numbers from speech. So, this association may additionally reflect that of a reduced capacity for activities requiring abstract processing from verbal command to motor action and limit engagement in functional ADLs.

The language domain features prominently. Here we found an observed association between depression and impairments in a picture naming task of common animals and objects. This finding aligns with the literature implicating aphasia as a risk factor for post-stroke depression symptomology.^39^ Additionally, retrieval of common objects may alternatively further index an association between depression symptomology and difficulties with verbal memory.

The current finding that non-neglect-specific performance accuracy was associated with increased depression symptomology aligns with the findings from Rock and colleagues’^40^ meta-analysis, indicating persistent deficits to attention in depression extra to the presence of low mood itself. Attentional mechanisms are composed of various factors, such as effort, sustained attention, and susceptibility to distraction – all of which can be reduced following stroke.^41^ Consequently, acquired impairments in these attentional factors following a stroke could potentially lessen the degree of positive reinforcement gained from activities, resulting in low mood. Moreover, the exertion required for daily activities post-stroke may contribute to attentional fatigue, potentially increasing the severity of depressive symptoms further. This is particularly relevant to the OCS visuospatial subtask, the broken hearts task, being both effortful and visually complex which may exacerbate the exertion element of spatial attention.

In contrast to Williams and Demeyere^15^ impairments in the motor imitation task – the single task comprising the praxis domain – only predicted depression severity when accounting for co-occurring anxiety severity. This increase in alpha (p) values counters the decrease expected when adding covariates in multiple regression models.^42^ Owing to the direct comparison of results between the Depression Only models and Depression with Anxiety models, this indicates potential collider bias of the anxiety severity variable, limiting internal validity for the praxis models.^43^ One explanation for the weaker association between praxis impairment and depression in a more moderately affected stroke sample could be that its functional impact may be less noticeable if moderate stoke leads to a reduction in activities requiring physical coordination.

### Clinical Implications and Future Research

Methodologically, the scope of these findings is limited to only those cognitive subcomponents as measured in the OCS. Thus, this is neither an exhaustive nor comprehensive list of all cognitive impairments that could potentially predict depression severity, such as speed of information processing, auditory attention, and visual perception difficulties.^44^ Rather, the novel associations identified between specific within-domain specific and depression severity highlight new pathways for researching strategies to augment post-stroke depression treatments. Impairments in these specific cognitive abilities may also have implications for those at risk of sustained depression severity and contribute to conceptual models of how depression is maintained following stroke.

The magnitude of the associations between cognitive impairments and depression severity without accounting for anxiety were relatively small.^45^ This was both corroborated by findings of the analysis of the pooled imputation data and by the findings of Williams and Demeyere^15^. Future replications should extend these results and consider combining cognitive impairment with other factors known to exacerbate low mood in stroke – such as fatigue, psychosocial isolation, and socioeconomic deprivation.^10,46^

### Limitations

Conceptually, the direction of causality between these identified associations is indeterminable without experimental evidence or longitudinal data. Outside of stroke, depression has been shown to induce cognitive changes, like reduced speed of processing.^44,47^ So, one could potentially argue that depression induces these cognitive impairments irrespective of stroke. Preliminary evidence suggests that acute post-stroke depression outcomes could predict longer-term post-stroke cognitive outcomes^48^ – yet this approach is confounded by the drastic mood change following acute stroke that do not reliably predict of longer-term mood-outcomes.^49^ Additionally, it is reasonable to assume some directional effect from impairment to depression because the OCS was designed to identify pronounced stroke-specific cognitive impairments, rather than population-level changes in cognition.

Statistically, mixing self-report measures (depression and anxiety severity) with objective measures (education and cognitive impairment) increases the risk of common method variance (CMV) bias.^50^ This means that variance explaining the outcome could be conflated with the unstandardised variance between these methods of outcome collection, reducing face validity. Presence of CMV bias is indicated by the increase in variance of depression severity each model explained after accounting for co-occurring anxiety severity. Given that self-reported depression and anxiety do frequently co-occur,^21^ CMV bias may also be additive in this context, because the OCS is not based on self-report measures. Analysing continuous cognitive-outcome data could increase sensitivity of findings, mitigating some CMV bias. The present analysis of binarised scores informed those thresholds of cognition specifically designed to determine functional impairments. However, a logical next step is a replication of this study with more sensitive, continuous cognitive measures to explore the association of subtask-specific cognitive ability – rather than impairment – with post-stroke depression.

## Conclusion

Domain-specific cognitive impairments can differentially influence the severity of post-stroke depressive symptoms. Specifically, impairments in calculation, episodic memory, picture naming, number writing, and non-neglect-specific visuospatial attention tasks can all predict post-stroke depression symptomology, even after controlling for co-occurring anxiety severity. Identifying these novel associations could help augment existing therapeutic interventions in light of these impairments, potentially enhancing therapeutic outcomes for those stroke survivors most at risk for depression.

## Data Availability

Data is freely available at https://www.dementiasplatform.uk/ and analysis code at https://osf.io/yrth4/

## Acknowledgements

The authors thank all participants and members of the Oxford Translational Neuropsychology Group for contributions to recruitment/testing.

