## Supplementary Material for "Specific Within-Domain Cognitive Impairments Predict Depression Severity Six-Months After Stroke"

**Supplementary Material Table of Contents**

**Supplementary Material A: Cohort Comparison**

This supplementary Material summarises the differences in stroke severity and demographics between the cohort analysed by Williams and Demeyere (2021) (referred to here as OCS-Care) and the cohort analysed in the present study (referred to here as OCS-Recovery). The present cohort was consecutively recruited from an acute stroke ward (*N* = 866) at a 6-month post-stroke assessment (N=430). Full details of reasons for attrition are reported in Milosevich et al. (2023). Figure A shows details a density plot of stroke severity and Table A.1 details the differences between cohorts in demographics. Table A.2 compares the prevalence of domain-specific cognitive impairment by cohort.

**Figure A**:

*National Institutes of Health Stroke Scale Scores by Oxford Cognitive Screen (OCS) Cohort*

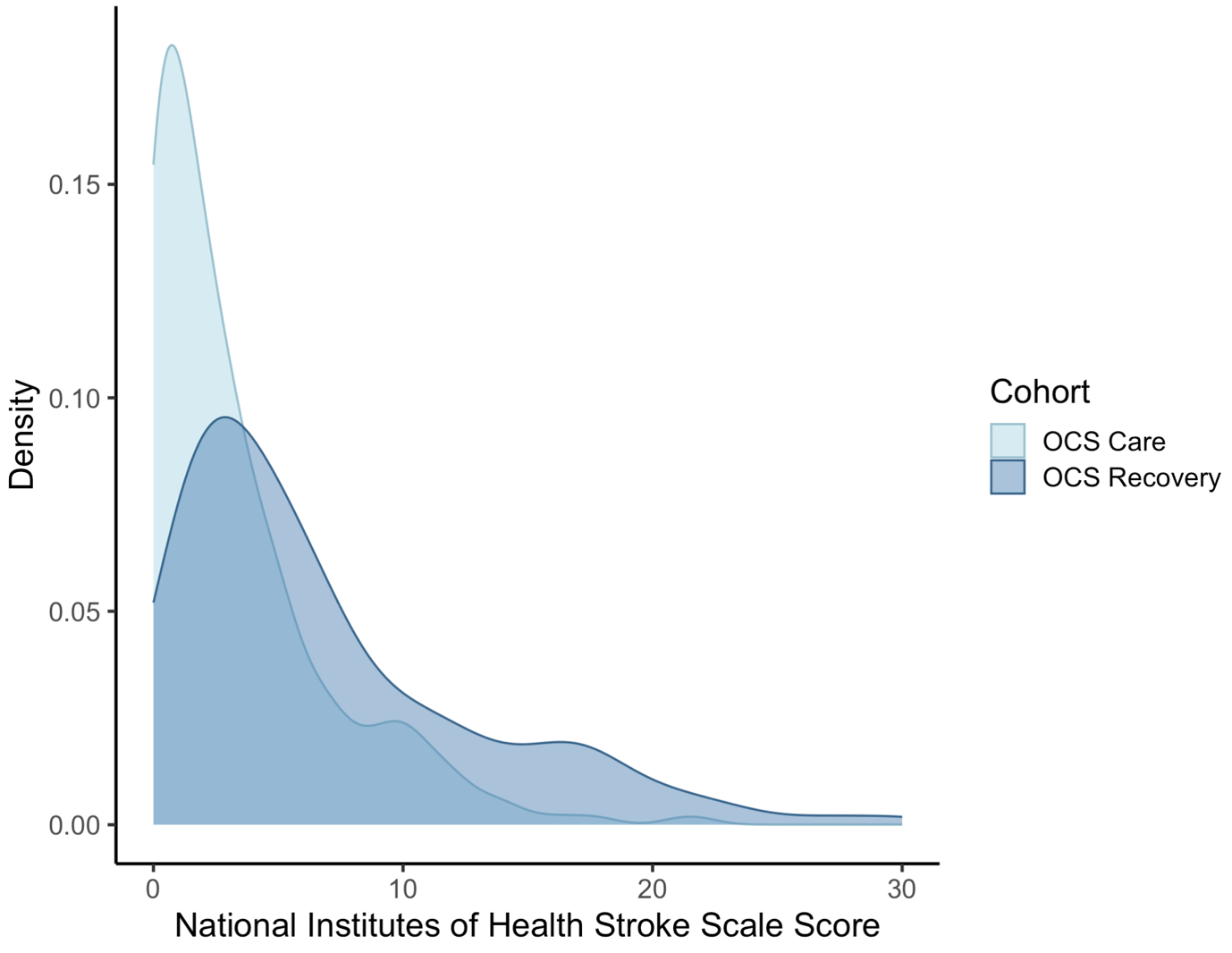

*Note.* This study analyses data from the OCS-Recovery cohort to replicate Williams and Demeyere’s (2021) analysis of the OCS-Care cohort.

**Table A.1**

*Comparison of Demographic, Mood, and Cognitive Impairment Data by OCS Cohort*

|  | Mean (*SD*) of Cohort | | *t* statistic (*df*) | *p* value |
| --- | --- | --- | --- | --- |
|  | OCS-Care | OCS-Recovery |  |  |
| Age (years) | 69.27 (12.12) | 73.86 (12.51) | -5.46 (863.34) | < .001* |
| HADS-D | 5.38 (4.06) | 5.43 (4.10) | -0.51 (805.08) | .610 |
| HADS-A | 5.76 (4.28) | 5.80 (4.35) | -0.12 (803.48) | .906 |
|  | Mean (*SD*) of Cohort | | *W* | *p* value |
| Education (years) | 11.82 (2.87) | 12.25 (3.55) | 91284 | .6183 |
| NIHSS | 3.32 (3.70) | 6.83 (6.01) | 39840 | <.001* |
|  | Percentage of Cohort (%) | | χ^2^ (*df*) | *p* value |
| Male Sex | 51.71 | 54.03 | 2.15 (1) | .143 |
| HADS-D **>** 7 | 26.32 | 26.23 | 0.15 (1) | .697 |
| HADS-A **>** 7 | 30.43 | 32.21 | 0.55 (1) | .457 |

*Note.* OCS = Oxford Cognitive Screen. This study analyses data from the OCS-Recovery cohort to replicate Williams and Demeyere’s (2021) analysis of the OCS-Care cohort.

HADS = Hospital Anxiety and Depression Scale where D and A are the depression and anxiety subscales and scores > 7 indicate ‘possible’ diagnostic depression/anxiety. NIHSS = National Institutes of Health Stroke Scale. Two tailed parametric *t*-tests compared groups for which the Levene’s tests were non-significant, Mann Whitney U tests (*W*) were used where Levene’s tests were significant, two-tailed Yates-corrected chi-squared tests were used for binary data. *Significant *p <* .050.

**Table A.2**

*Prevalence of Domain-Specific Impairments on the Oxford Cognitive Screen (OCS) by Cohort*

| Cognitive Domain | Impairment Prevalence (%) | | χ^2^ (*df*) | *p* value |
| --- | --- | --- | --- | --- |
|  | OCS-Care | OCS-Recovery |  |  |
| Language | 17.85 | 32.63 | 425.46 (1) | < .001* |
| Memory | 18.99 | 32.32 | 425.44 (1) | < .001* |
| Spatial Attention | 29.50 | 34.91 | 425.96 (1) | < .001* |
| Executive Function | 10.76 | 24.40 | 367.6 (1) | < .001* |
| Numeracy | 15.79 | 23.06 | 424.34 (1) | < .001* |
| Praxis | 9.84 | 19.80 | 397.73 (1) | < .001* |

*Note.* Two-tailed Yates-corrected chi-squared tests were performed on raw frequency counts, not percentages. This study analyses data from the OCS-Recovery cohort to replicate Williams and Demeyere’s (2021) analysis of the OCS-Care cohort.

**Supplementary Material B: Anxiety Analysis**

This is a replication of Williams and Demeyere (2021) with the present cohort, investigating the domain-general and domain-specific cognitive impairments n the Oxford Cognitive Screen that predict anxiety severity. This was done both before and after covarying for co-occurring depression severity, in the ‘Anxiety Only’ and ‘Anxiety with Depression’, models, respectively.

**Association of Cognitive Impairment with Anxiety**

***Anxiety Model Covariates***

HADS-A significantly correlated with HADS-D (*r*_(383)_ = .63 *p* < .001), lower age at time of stroke (*r*_(383)_ = -.13 *p* < .001), and sex (*t*_(383)_ = -.11 *p* = .034). Therefore, age and sex were be included as covariates in all models predicting anxiety severity. Depression will be included as a covariate for Anxiety with Depression Models only.

1. ***Domain-General Models Predicting Anxiety Severity***

**Anxiety Only Model.**

There was a significant model predicting depression severity by domain-general impairment, *F*_(3, 381)_ = 7.60, *adjusted R^2^* = .05, *p*  < .001 before controlling for co-occurring depression severity. Total number of concurrent impairments was a significant predictor in the model (*b*_(.85)_ = 2.62, *t* = 3.07, 95% CI [0.94, 4.30], *p* = .002), as well as age (*b*_(.02)_ = -0.07, *t* = -3.64, 95% CI [-0.10, -0.03], *p* < .001), and sex (*b*_(.44)_ = 1.07, *t* = 2.45, 95% CI [0.21, 1.93], *p* = .015).

**Anxiety Only Model.**

After controlling for co-occurring depression severity, total number of concurrent impairments no longer predicted anxiety severity (*b*_(.69)_ = -0.06, *t* = -0.09, 95% CI [-1.41, 1.29], *p* = .928) and was instead predicted by age (*b*_(.01)_ = -0.06, *t* = -3.95, 95% CI [-0.08, -0.03], *p* < .001), sex (*b*_(.34)_ = 0.95, *t* = 2.78, 95% CI [0.28, 1.62], *p* = .006), and depression severity (*b*_(.04)_ = 0.67, *t* = 15.81, 95% CI [0.59, 0.75], *p* < .001).

2. ***Domain-Specific Models Predicting Anxiety Severity***

**Anxiety Only Models.**

Without accounting for co-occurring depression severity, only language (*b*_(.48)_ = 1.17, *t* = 2.46, 95% CI [0.24, 2.11], *p* = .011), memory (*b*_(.48)_ = 0.98, *t* = 2.05, 95% CI [0.04, 1.93], *p* = .041), and spatial attention (*b*_(.45)_ = 1.24, *t* = 2.73, 95% CI [0.35, 2.12], *p* = .007) predicted anxiety severity. Neither numeracy (*p* = .052), praxis (*p* = .454), nor executive function (*p* = .586) were significant predictor variables. Pooled imputation results aligned with complete case analysis for all domain-specific impairments (complete case *t*-values range = 0.54 [executive function] ­– 2.73 [spatial attention]; pooled imputation *t*-values range = 0.24 [executive function] – 2.87 [spatial attention]: Appendix Imputation),

Age was a significant predictor in all models (*p* range = .001 [spatial attention] – .027 [executive function]) with a negative association with HADS-A. Sex was a significant predictor in all models (*p* range = .010 [language] – .018 [numeracy]) except for executive function (*p* = .084) where male sex shared a positive association with HADS-A.

Effect sizes were small for all models (*Adjusted R^2^* range = .03 [praxis] – .04 [all other models]).

**Anxiety with Depression Models.**

After accounting for co-occurring depression severity, no domain-specific cognitive impairments variables were significant predictor variables in complete cases (*p* range = .429 – .937). Pooled imputation results aligned with complete case analysis for all domain-specific impairments. Age was a significant predictor in all models (*p*s ≤ .001) sharing a negative association with HADS-A. Sex (*p* range = .005 – .020) and HADS-D scores (*ps* < .001) were significant predictors in all models sharing a positive association with HADS-A scores.

All overall models predicting anxiety severity by cognitive impairment were significant (*p* < .001) and effect sizes were large (*Adjusted R^2^* range = .40 [executive function] – .43 [language, numeracy, praxis, and spatial attention]).

***Subtask-Specific Models Predicting Anxiety Severity***:

Subtask specific models were not explored because no domain-general or domain-specific post-stroke cognitive impairments predicted anxiety severity after covarying for co-occurring depression severity.

**Supplementary Material C: Pooled Imputations**

This supplementary material details the analyses of data from pooled imputations. Five data sets were generated using predictive mean matching. As per the analysis plan, multiple regression analysis was performed on each of these five imputed data individually. Then, pooled statistics of these were used as the pooled model statistics. This was to maximise use of data (*N* = 430) to compare with complete case data (*n* = 385) for stability of findings.

1. The pooled model statistics the *domain-specific* cognitive impairment variables or both the Depression Only Models and the Depression with Anxiety Models are presented in Table C.1.
2. The pooled model statistics the *subtask-specific* cognitive impairment variables or both the Depression Only Models and the Depression with Anxiety Models are presented in Table C.2.

**Table C.1**:

*Domain-Specific OCS Impairment Predictor Variable Statistics for the 12 Models Predicting Depression Severity using Pooled Imputation Data, With and Without Anxiety Severity.*

|  | Impairment Predictor Variable Statistics | | | | | | |
| --- | --- | --- | --- | --- | --- | --- | --- |
|  | Depression Only Model | | |  | Depression with Anxiety Model | | |
| Impairment | *b_1_* (*SE*) | *t* statistic | *p* value |  | *b_1_* (*SE*) | *t* statistic | *p* value |
| Language | 1.34 (.45) | 2.98 | .003* |  | .81 (.34) | 2.41 | .016* |
| Numeracy | 1.98 (.56) | 3.57 | <.001 * |  | 1.31 (.40) | 3.31 | .001* |
| Praxis | 1.00 (.52) | 1.92 | .056 |  | 1.03 (.43) | 2.40 | .019* |
| Memory | 1.57 (.53) | 2.95 | .006* |  | 1.03 (.36) | 2.89 | .004* |
| Attention | 1.42 (.43) | 3.32 | .001* |  | .94 (.32) | 2.92 | .004* |
| Executive Function | .78 (.52) | 1.50 | .137 |  | .83 (.41) | 2.00 | .051 |

*Note.* The Depression Only model formula is Depression Severity ~ *b_0_ + b_1_*Impairment + *b_2_*Education + ϵ. The formula for the Depression with Anxiety Model is Depression Severity ~ *b_0_ + b_1_*Impairment + *b_2_*Education + *b_3_*Anxiety Severityϵ.

**Table C.2**:

*Subtask-Specific OCS Impairment Predictor Variable Statistics for the 12 Models Predicting Depression Severity using Pooled Imputation Data, With and Without Anxiety Severity.*

|  | Impairment Predictor Variable Statistics | | | | | | |
| --- | --- | --- | --- | --- | --- | --- | --- |
|  | Depression Only Model | | |  | Depression with Anxiety Model | | |
| Impairment | *b_1_* (*SE*) | *t* statistic | *p* value |  | *b_1_* (*SE*) | *t* statistic | *p* value |
| Picture Naming | 1.31 (.64) | 2.04 | .051 |  | 1.29 (.53) | 2.44 | .024* |
| Semantic Task | 3.73 (1.28) | 2.91 | .022* |  | 2.14 (1.21) | 1.78 | .127 |
| Sentence Reading | .85 (.56) | 1.51 | .138 |  | .52 (.47) | 1.10 | .280 |
| Number Writing | 1.73 (.56) | 3.12 | .003* |  | 1.22 (.44) | 2.75 | .008* |
| Calculation | 2.74 (.67) | 4.00 | < .001* |  | 1.64 (.55) | 2.97 | .004* |
| Orientation | 1.05 (.66) | 1.58 | .126 |  | 1.00 (.52) | 1.94 | .064 |
| Verbal Memory | .73 (.55) | 1.33 | .185 |  | .56 (.44) | 1.26 | .211 |
| Episodic Memory | 2.01 (.61) | 3.34 | < .001* |  | 1.41 (.48) | 2.96 | .003* |
| Broken Heart Task | 1.62 (.48) | 3.37 | .001* |  | 1.28 (.35) | 3.71 | <.001* |
| Space Attention | .46 (.68) | .68 | .497 |  | .72 (.51) | 1.43 | .157 |
| Object Attention | .50 (.54) | .92 | .364 |  | .14 (.38) | .36 | .716 |

*Note.* The Depression Only model formula is Depression Severity ~ *b_0_ + b_1_*Impairment + *b_2_*Education + ϵ. The formula for the Depression with Anxiety Model is Depression Severity ~ *b_0_ + b_1_*Impairment + *b_2_*Education + *b_3_*Anxiety Severityϵ.

**Supplementary Material D: Depression Only Model Statistics**

This supplementary material details the model statistics for the ‘Depression Only’ models using complete case data predicting depression severity by domain-specific and subtask-specific impairment on the Oxford Cognitive Screen (OCS).

1. First, Table D.1 summarises the model statistics for the six models predicting depression severity.
2. Then, Table D.2 summarises the statistics for the individual predictor variables within these models.
3. Then, Table D.3 does something slightly different. This table summarises individual predictor variables within the Depression Only models but for the subtask-specific cognitive impairments on the OCS that predict depression severity. The overall model statistics are summarised in text.

**Table D.1**:

*Model Statistics of The Six ‘Depression Only Models’ Predicting Depression Severity by Domain-Specific OCS Cognitive Impairments and Education*

|  | Depression Severity ~ *b_0_ + b_1_* Impairment + *b_2_* Education + ϵ | | |
| --- | --- | --- | --- |
| Impairment | *F* statistic (*df*) | *AdjR^2^* | *p* value |
| Language | 5.73 (2, 379) | .02 | .021* |
| Numeracy | 8.32 (2, 379) | .04 | .002* |
| Praxis | 3.55 (2, 363) | .01 | .179 |
| Memory | 6.40 (2, 379) | .03 | .011* |
| Executive Function | 3.28 (2, 332) | .01 | .232 |
| Spatial Attention | 6.59 (2, 379) | .03 | .009* |

*Note.* This table summarises six separate models predicting depression severity by each of the six domain-specific impairments on the Oxford Cognitive Screen (OCS), covarying for education. *Significant (*p* < .050) after false discovery rate (FDR) correction for multiple comparisons.

**Table D.2**:

*Predictor Variables of The Six ‘Depression Only Models’ Predicting Depression Severity by Domain-Specific OCS Cognitive Impairments and Education*

|  |  | Predictor Variable Statistics | | | |
| --- | --- | --- | --- | --- | --- |
|  | Predictor Variable | *b (SE)* | *t* statistic | 95% CI | *p* value |
| Impairment | Language | 1.24 (.46) | 2.70 | [0.33, 2.14] | .007* |
|  | Education | -.09 (.06) | -1.52 | [-0.02, 0.02] | .129 |
| Impairment | Numeracy | 1.86 (.53) | 3.52 | [0.82, 2.89] | <.001* |
|  | Education | -.08 (.06) | -1.36 | [-0.19, 0.04] | .175 |
| Impairment | Praxis | .95 (.53) | 1.78 | [-0.10 2.00] | .077 |
|  | Education | -.11 (.06) | -1.91 | [-0.23, 0.01] | .056 |
| Impairment | Memory | 1.35 (.46) | 2.93 | [0.45, 2.26] | .004* |
|  | Education | -.09 (.06) | -1.52 | [-0.20, 0.03] | .128 |
| Impairment | Executive Function | .80 (.51) | 1.56 | [-0.21, 1.82] | .120 |
|  | Education | -.12 (.06) | -1.85 | [-0.24, 0.01] | .065 |
| Impairment | Spatial Attention | 1.27 (.42) | 3.00 | [0.44, 2.11] | .003* |
|  | Education | -.09 (.06) | -1.49 | [-0.20, 0.02] | .138 |

*Note.* *Significant (*p* <.050) after FDR correction for multiple tests. HADS-D = Hospital Anxiety and Depression Scale – Depression Subscale

**Table D.3**:

*Predictor Variables in the ‘Depression Only Models’ Predicting Depression Severity by Subtask-Specific OCS Cognitive Impairments and Education*

|  |  | Predictor Variable Statistics | | | |
| --- | --- | --- | --- | --- | --- |
|  | Predictor Variable | *b (SE)* | *t* statistic | *95% CI* | *p* value |
| Impairment | Picture Naming | 1.09 (.57) | 1.91 | [-0.03, 2.22] | .057 |
|  | Education | -.09 (.06) | -1.59 | [-0.21, 0.02] | .112 |
| Impairment | Semantic Task | 3.18 (1.25) | 2.53 | [0.70, 5.65] | .012* |
|  | Education | -.11 (.07) | -1.52 | [-0.24, 0.03] | .129 |
| Impairment | Sentence Reading | .88 (.54) | 1.64 | [-0.17, 1.93] | .101 |
|  | Education | -.10 (.06) | -1.78 | [-0.22, 0.01] | .076 |
| Impairment | Number Writing | 1.58 (.58) | 2.71 | [0.43, 2.73] | .007* |
|  | Education | -.08 (.06) | -1.35 | [-0.20, 0.04] | .177 |
| Impairment | Calculation | 2.48 (.72) | 3.44 | [1.06, 3.89] | < .001* |
|  | Education | -.08 (.06) | -1.44 | [-0.20, 0.03] | .149 |
| Impairment | Orientation | .71 (.58) | 1.23 | [-0.43, 1.86] | .218 |
|  | Education | -.10 (.06) | -1.75 | [-0.22, 0.01] | .080 |
| Impairment | Verbal Memory | .77 (.56) | 1.38 | [-0.33, 1.87] | .168 |
|  | Education | -.11 (.06) | -1.88 | [-0.23, 0.01] | .060 |
| Impairment | Episodic Memory | 2.03 (.65) | 3.11 | [0.75, 3.31] | .002* |
|  | Education | -.09 (.06) | -1.62 | [-0.21, 0.02] | .107 |
| Impairment | Broken Heart Task | 1.52 (.45) | 3.39 | [0.64, 2.40] | < .001* |
|  | Education | -.09 (.06) | -1.52 | [-0.20, 0.03] | .130 |
| Impairment | Space Attention | .54 (.64) | .85 | [-0.71, 1.79] | .397 |
|  | Education | -.11 (.06) | -1.94 | [-0.23, 0.01] | .053 |
| Impairment | Object Attention | .45 (.50) | .89 | [-0.54, 1.43] | .374 |
|  | Education | -.12(.06) | -1.98 | [-0.23, -0.01] | .048 |

*Note.* *Significant (*p* <.050) after FDR correction for multiple tests. HADS-D = Hospital Anxiety and Depression Scale – Depression Subscale

**Supplementary Material E: Depression with Anxiety Model Statistics**

This supplementary Material details the model statistics for the ‘Depression with Anxiety’ models using complete case data predicting depression severity by domain-specific and subtask-specific impairment on the Oxford Cognitive Screen (OCS).

1. First, Table E.1 summarises the model statistics for the six models predicting depression severity.
2. Then, Table E.2 summarises the statistics for the individual predictor variables within these models (OCS domain-specific impairment, education, and anxiety).
3. Then, Table E.3 does something slightly different. This table summarises individual predictor variables within the ‘Depression with Anxiety’ models but for the subtask-specific cognitive impairments on the OCS that predict depression severity. The overall model statistics are summarised in text.

**Table E.1**:

*Model Statistics of The Six ‘Depression with Anxiety Models’ Predicting Depression Severity by Domain-Specific OCS Cognitive Impairments, Education, and Anxiety Severity*

|  | Depression Severity ~ *b_0_ + b_1_* Impairment + *b_2_* Education + *b_3_* Anxiety Severity + ϵ | | |
| --- | --- | --- | --- |
| Impairment | *F* statistic (*df*) | *AdjR^2^* | *p* value |
| Language | 86.08 (3, 378) | .40 | < .001* |
| Numeracy | 88.87 (3, 378) | .41 | < .001* |
| Praxis | 82.67 (3, 361) | .40 | < .001* |
| Memory | 87.13 (3, 378) | .40 | < .001* |
| Executive Function | 70.15 (3, 331) | .38 | < .001* |
| Spatial Attention | 87.19 (3, 378) | .40 | < .001* |

*Note.* This table summarises six separate models predicting depression severity by each of the six domain-specific impairments on the Oxford Cognitive Screen (OCS), as well as anxiety severity, and education. *Significant (p < .050) after false discovery rate (FDR) correction for multiple comparisons.

**Table E.2**:

*Predictor Variables of The Six ‘Depression with Anxiety Models’ Predicting Depression Severity by Domain-Specific OCS Cognitive Impairments, Education, and Anxiety Severity*

|  |  | Predictor Variable Statistics | | | |
| --- | --- | --- | --- | --- | --- |
|  | Predictor Variable | *b (SE)* | *t* statistic | 95% CI | *p* value |
| Impairment | Language | .76 (.36) | 2.11 | [0.05, 1.47] | .036* |
|  | Education | -.04 (.05) | -.76 | [-0.12, 0.06] | .447 |
|  | Anxiety | 0.58 (.04) | 15.48 | [0.51, 0.66] | < .001* |
| Impairment | Numeracy | 1.28 (.42) | 3.01 | [0.46, 2.10] | .002* |
|  | Education | -.03 (.05) | -.58 | [-0.12, 0.06] | .564 |
|  | Anxiety | 0.58 (.04) | 15.48 | [0.51, 0.66] | < .001* |
| Impairment | Praxis | .94 (.42) | 2.27 | [0.89, 3.48] | .024* |
|  | Education | -.04 (.05) | -.96 | [-0.13, 0.06] | .339 |
|  | Anxiety | 0.59 (.04) | 15.37 | [0.51, 0.66] | < .001* |
| Impairment | Memory | .91 (.36) | 2.52 | [0.20, 1.63] | .012* |
|  | Education | -.03 (.05) | -.73 | [-0.12, 0.06] | .469 |
|  | Anxiety | 0.58 (.04) | 15.51 | [0.51, 0.66] | < .001* |
| Impairment | Executive Function | .75 (.41) | 1.82 | [-0.06, 1.55] | .068 |
|  | Education | -.04 (.05) | -.87 | [-0.14, 0.06] | .387 |
|  | Anxiety | 0.57 (.04) | 14.14 | [0.49, 0.64] | < .001* |
| Impairment | Spatial Attention | .85 (.33) | 2.54 | [0.19, 1.50] | .012* |
|  | Education | -.03 (.05) | -.70 | [-0.12, 0.06] | .483 |
|  | Anxiety | 0.58 (.04) | 15.49 | [0.51, 0.66] | < .001* |

*Note.* *Significant (*p* <.050) after FDR correction for multiple tests. HADS-D = Hospital Anxiety and Depression Scale – Depression Subscale

**Table E.3**:

*Predictor Variables of The Six ‘Depression with Anxiety Models’ Predicting Depression Severity by Subtask-Specific OCS Cognitive Impairments, Education, and Anxiety Severity*

|  |  | Predictor Variable Statistics | | | |
| --- | --- | --- | --- | --- | --- |
|  | Predictor Variable | *b (SE)* | *t* statistic | 95% CI | *p* value |
| Impairment | Picture Naming | 1.18 (.45) | 2.66 | [0.31, 2.06] | .008* |
|  | Education | -.03 (.05) | -0.61 | [-0.12, 0.06] | .540 |
|  | Anxiety | .59 (.04) | 15.73 | [0.52, 0.66] | < .001* |
| Impairment | Semantic Task | 1.37 (1.01) | 1.35 | [-0.63, 3.36] | .179 |
|  | Education | -.04 (.05) | -0.78 | [-0.15, 0.07] | .437 |
|  | Anxiety | .55 (.05) | 11.44 | [0.46, 0.65] | < .001* |
| Impairment | Sentence Reading | .47 (.42) | 1.13 | [-0.35, 1.30] | .260 |
|  | Education | -.05 (.05) | -0.99 | [-0.13, 0.05] | .323 |
|  | Anxiety | .58 (.04) | 15.37 | [0.51, 0.66] | < .001* |
| Impairment | Number Writing | 1.17 (.46) | 2.54 | [0.26, 2.07] | .011* |
|  | Education | -.03 (.05) | -0.64 - | [-0.12, 0.06] | .520 |
|  | Anxiety | .58 (.04) | 15.42 | [0.51, 0.66] | < .001* |
| Impairment | Calculation | 1.44 (.57) | 2.51 | [0.31, 2.56] | .012* |
|  | Education | -.04 (.05) | 0.80 | [-0.13, 0.05] | .423 |
|  | Anxiety | .58 (.04) | 15.06 | [0.50, 0.65] | < .001* |
| Impairment | Orientation | .75 (.45) | 1.66 | [-0.14, 1.64] | .098 |
|  | Education | -.04 (.05) | -0.86 | [-0.13, 0.05] | .391 |
|  | Anxiety | .59 (.04) | 15.71 | [0.52, 0.67] | < .001* |
| Impairment | Verbal Memory | .56 (.44) | 1.28 | [-0.30, 1.42] | .203 |
|  | Education | -.04 (.05) | -0.95 | [-0.14, 0.05] | .345 |
|  | Anxiety | .58 (.04) | 15.18 | [0.50, 0.65] | < .001* |
| Impairment | Episodic Memory | 1.36 (.52) | 2.63 | [0.34, 2.37] | .009* |
|  | Education | -.03 (.05) | -0.74 | [-0.13, 0.06] | .458 |
|  | Anxiety | .57 (.04) | 15.06 | [0.50, 0.65] | < .001* |
| Impairment | Broken Heart Task | 1.24 (.35) | 3.50 | [0.54, 1.93] | <.001* |
|  | Education | -.03 (.05) | -0.60 | [-0.12, 0.06] | .547 |
|  | Anxiety | .58 (.04) | 15.23 | [0.50, 0.65] | < .001* |
| Impairment | Space Attention | .75 (.50) | 1.51 | [-0.23, 1.73] | .132 |
|  | Education | -.04 (.05) | -0.90 | [-0.13, 0.05] | .367 |
|  | Anxiety | .59 (.04) | 15.27 | [0.51, 0.65] | < .001* |
| Impairment | Object Attention | .14 (.38) | .20 | [-0.70, 0.86] | .716 |
|  | Education | -.05 (.05) | -1.06 | [-0.14, 0.04] | .290 |
|  | Anxiety | .59 (.04) | 15.14 | [0.51, 0.66] | < .001* |

*Note.* *Significant (*p* <.050) after FDR correction for multiple tests. HADS-D = Hospital Anxiety and Depression Scale – Depression Subscale

**Supplementary Material F: Univariate Models**

This supplementary material details the output of the univariate models predicting depression severity by domain-specific task impairments (Table F.1) and subtask-specific impairments (Table F.2) on the OCS. This was done to create the visual heatmap of associations overlaid on the OCS wheel in Figure 2 in text. The statistics in this table were solely used to populate the heatmap on the OCS wheel in Figure 2 in text to show the strength of association between depression severity and with these impairments without covariates.

**Table F.1**:

*Model Statistics for the Six Univariate Models Predicting Depression Severity by Domain-Specific Cognitive Impairments on the Oxford Cognitive Screen.*

|  | Depression Severity ~ *b_0_ + b_1_* Impairment + ϵ | | | | | | | |
| --- | --- | --- | --- | --- | --- | --- | --- | --- |
|  | Overall Regression Model | | |  | Impairment Predictor Variable | | | |
| Impairment | *F* statistic (*df*) | *AdjR^2^* | *p* value |  | *b_1_* (*SE*) | *t* statistic | 95% CI | *p* value |
| Language | 9.04 (1, 383) | .02 | .017* |  | 1.35 (.45) | 3.01 | [0.47, 2.23] | .003* |
| Numeracy | 14.19 (1, 383) | .03 | .001* |  | 1.93 (.51) | 3.77 | [0.92, 2.94] | <.001* |
| Praxis | 3.36 (1, 366) | .01 | .406 |  | 0.98 (.53) | 1.83 | [-0.07, 2.03] | .068 |
| Memory | 10.10 (1, 38) | .02 | .010* |  | 1.44 (.45) | 3.18 | [0.55, 2.33] | .002* |
| Executive Function | 2.97 (1, 335) | .01 | .513* |  | 0.88 (.51) | 1.72 | [-0.12, 1.89] | .086* |
| Spatial Attention | 11.21 (1, 383) | .03 | .005* |  | 1.39 (.42) | 3.35 | [0.57, 2.21] | .001* |

*Note.* *Significant (p < .050) after false discovery rate (FDR) correction for multiple comparisons.

**Table F.2**:

*Model Statistics for the 11 Univariate Models Predicting Depression Severity by Domain-Specific Cognitive Impairments on the Oxford Cognitive Screen.*

|  | Depression Severity ~ *b_0_ + b_1_* Impairment + ϵ | | | | | | | |
| --- | --- | --- | --- | --- | --- | --- | --- | --- |
|  | Overall Regression Model | | |  | Impairment Predictor Variable | | | |
| Impairment | *F* statistic (*df*) | *AdjR^2^* | *p* value |  | *b_1_* (*SE*) | *t* statistic | 95% CI | *p* value |
| Picture Naming | 4.97 (1, 382) | .01 | .290 |  | 1.25 (0.56) | 2.23 | [0.15, 2.36] | .026* |
| Semantic Task | 6.18 (1, 230) | .02 | .150 |  | 3.12 (1.26) | 2.49 | [0.65, 5.60] | .014* |
| Sentence Reading | 3.55 (1, 380) | .01 | .662 |  | 1.00 (0.53) | 1.89 | [-0.04, 2.04] | .060 |
| Number Writing | 8.51 (1, 377) | .02 | .041 |  | 1.65 (0.57) | 2.92 | [0.54, 2.77] | .004* |
| Calculation | 13.32 (1, 376) | .03 | .003 |  | 2.61 (0.71) | 3.65 | [1.20, 4.01] | <.001* |
| Orientation | 2.47 (1, 381) | <.01 | 1.00 |  | 0.90 (0.57) | 1.57 | [-0.22, 2.02] | .117 |
| Verbal Memory | 2.49 (1, 374) | <.01 | 1.00 |  | 0.88 (0.56) | 1.58 | [-0.22, 1.97] | .115 |
| Episodic Memory | 10.11 (1, 374) | .02 | .018 |  | 2.04 (0.64) | 3.18 | [0.78, 3.30] | .002* |
| Broken Heart Task | 14.14 (1, 376) | .03 | .002 |  | 1.65 (0.44) | 3.76 | [0.79, 2.51] | <.001* |
| Space Attention | 1.11 (1, 372) | <.01 | 1.00 |  | 0.67 (0.63) | 1.06 | [-0.58, 1.91] | .292 |
| Object Attention | 1.17 (1, 372) | <.01 | 1.00 |  | 0.54 (0.50) | 1.08 | [-0.44, 1.52] | .279 |

*Note.* *Significant (p < .050) after false discovery rate (FDR) correction for multiple comparisons.
